# Outcomes of Atrial Fibrillation Catheter Ablation in Patients with Peripheral Artery Disease: A Nationwide Inpatient Sample Study

**DOI:** 10.64898/2026.05.22.26353913

**Authors:** Valentine C. Nriagu, Soroush Shakeri, Tagbo Charles Nduka, Paul-Alex Ifeagwazi, Aniekeme Etuk, Steven Sorci, Gregory Cunn, Rohan Patel, Shiv Raj, Jacob Shani, Esosa Odigie-Okon

## Abstract

**Background:** Peripheral artery disease (PAD) may amplify procedural risk during atrial fibrillation (AF) catheter ablation, but dedicated evidence is lacking.

We aimed to evaluate the association between PAD and in-hospital outcomes among adults undergoing AF ablation in the National Inpatient Sample (NIS).

**Methods:** We identified inpatient AF ablation hospitalizations in the 2016 through 2020 National Inpatient Sample using ICD-10-PCS procedure codes and a concurrent AF diagnosis. PAD was identified from ICD-10-CM diagnosis codes used in prior claims-based PAD studies. Stabilized inverse probability of treatment weighting based on the propensity score was used to balance baseline differences. The primary outcome was in-hospital mortality. Fourteen secondary outcomes and 2 composite end points were prespecified.

**Results:** Among 22,166 AF ablation hospitalizations, 899 (4.06%) involved patients with PAD. Compared with patients without PAD, those with PAD were older and had a substantially greater cardiovascular, renal, and smoking/tobacco comorbidity burden. In-hospital mortality did not differ significantly (1.39% vs 1.06%; aOR, 1.32; 95% CI, 0.66 - 2.64; P= 0.44). PAD was associated with higher odds of major bleeding (aOR, 1.62; 95% CI, 1.17 - 2.24; P = 0.004), vascular or access-site complications (aOR, 1.80; 95% CI, 1.04 - 3.12; P = 0.04), acute kidney injury (aOR, 1.31; 95% CI, 1.05 - 1.64; P = 0.02), and composite major adverse hospital events (aOR, 1.29; 95% CI, 1.05 - 1.59; P = 0.02). Total hospital charges were 13% higher (charge ratio, 1.13; 95% CI, 1.04 - 1.22; P = 0.003). Major bleeding, vascular/access-site complications, cardiac arrest, and composite major adverse in-hospital events remained elevated in sensitivity analysis. Conclusion. PAD was independently associated with higher bleeding risk, vascular or access-site complications, acute kidney injury, and composite major adverse hospital event during AF ablation, identifying a clinically relevant subgroup with elevated periprocedural risk.

**Clinical Perspective:** *What Is New?:*

- In a national inpatient cohort of adults undergoing atrial fibrillation catheter ablation, peripheral artery disease was associated with higher odds of major bleeding and vascular/access-site complications.
- Acute kidney injury and composite major adverse hospital events were higher in the primary weighted analysis, while in-hospital mortality was not significantly different.

*What Are the Clinical Implications?:*

- Peripheral artery disease may identify a higher-risk subgroup who may benefit from individualized antithrombotic planning, renal-protective strategies, and careful vascular access planning.

## INTRODUCTION

Atrial fibrillation (AF) is the most common sustained cardiac arrhythmia worldwide, affecting an estimated 59 million individuals globally, with prevalence projected to nearly double over the coming decades.^1,2^ AF is independently associated with a 5-fold increase in stroke risk, a 3-fold increase in heart failure, and significant excess mortality.^1,3^ The CHA₂DS₂-VASc score, widely used to stratify thromboembolic risk in AF, explicitly incorporates vascular disease, including peripheral artery disease (PAD), as a risk-modifying component, reflecting the recognized interplay between AF and systemic atherosclerosis.^4^

Catheter ablation has fundamentally reshaped the management of AF, evolving from a therapy reserved for drug-refractory patients to a guideline-endorsed first-line rhythm-control strategy.^3,5^ Landmark trials have demonstrated benefits of early rhythm control and catheter ablation in reducing cardiovascular endpoints and heart failure hospitalizations.^6–8^ However, ablation remains an invasive procedure, with a pooled major complication rate of approximately 4.5% across 89 randomized trials, dominated by cardiac tamponade, vascular access-site injury, and stroke.^9^ Newer energy modalities such as pulsed field ablation have shown promising early safety profiles,^10–12^ though their impact in high-risk subgroups remains to be defined.

PAD affects more than 230 million people globally and is recognized as a manifestation of systemic atherosclerosis and a marker of high cardiovascular risk.^13–16^ In contemporary AF registries, concomitant PAD is associated with higher rates of stroke, major bleeding, and mortality.^17,18^ The co-existence of AF and PAD is increasingly common in aging populations, yet the impact of PAD on periprocedural ablation safety has not been specifically evaluated.

This gap matters because transfemoral venous access, the dominant route for AF ablation, may be more hazardous in patients with aortoiliac calcification, where altered anatomy and vascular calcification can increase the risk of difficult access and access-site injury. PAD patients also carry heavier burdens of atherosclerotic cardiovascular disease, chronic kidney disease, heart failure, and coagulopathy, and they are often treated with antiplatelet therapy after peripheral revascularization.^19^ Together, these factors may amplify bleeding risk during ablation and increase renal vulnerability. Direct procedural evidence, however, remains limited. In one single-center study of AF ablation, peripheral vascular disease was associated with early cerebral thromboembolic complications, but contemporary data on bleeding, vascular complications, acute kidney injury, and resource use in patients with PAD are lacking.^20^ Prior inpatient analyses have examined sex, hypertrophic cardiomyopathy, and malignancy as modifiers of ablation outcomes,^21,22^ but PAD has not been evaluated as a dedicated exposure in a nationally representative US cohort.

Our study aimed to evaluate the association between concomitant PAD and in-hospital outcomes among adults undergoing AF catheter ablation, using nationally representative data. We hypothesized that PAD would be independently associated with increased adverse outcomes among AF patients undergoing catheter ablation.

## METHODS

### Data Source

We performed a retrospective cross-sectional study using the Healthcare Cost and Utilization Project National Inpatient Sample (NIS) from 2016 through 2020. The NIS is the largest all-payer inpatient database in the United States and contains discharge-level data from approximately 7 million hospitalizations annually, representing a 20% stratified sample of nonfederal acute care hospital discharges. Diagnoses are coded using ICD-10-CM (International Classification of Diseases, Tenth Revision, Clinical Modification) and procedures using ICD-10-PCS (Procedure Coding System). We included adults aged 18 years or older.

The study was exempt from institutional review board approval because the NIS contains only de-identified discharge data.

### Cohort Identification

We identified AF ablation hospitalizations using ICD-10-PCS procedure codes previously used in administrative analyses of inpatient pulmonary vein isolation and AF ablation. Because contemporary ICD-10-PCS codes are imperfect for identifying pulmonary vein isolation when used in isolation, as shown by Ferro et al,^23^ we required a concurrent AF diagnosis and applied additional exclusions to improve specificity.

Specifically, hospitalizations carrying code 02583ZZ together with permanent pacemaker implantation during the index admission were excluded because that combination more likely reflects atrioventricular junction ablation than pulmonary vein isolation.

We required a concurrent ICD-10-CM diagnosis of AF for all included procedure codes to improve capture of AF-directed ablation. AF was confirmed by at least 1 validated ICD-10-CM diagnosis code. Hospitalizations coded for atrial flutter without AF were excluded.

All diagnosis and procedure codes are provided in the Appendix.

### Exposure Definition: Peripheral Artery Disease

The exposure of interest was peripheral artery disease (PAD) documented as a comorbid diagnosis during the AF ablation hospitalization. PAD was identified using an ICD-10-CM code set drawn from prior claims-based PAD studies. ^24^ Hospitalizations with any qualifying PAD code in any diagnosis position were classified as PAD-exposed.

### Outcome Definitions

#### Primary Outcome

The primary outcome was in-hospital mortality which was analyzed without further modification.

### Secondary Outcomes

We included pre-specified secondary outcomes each identified from validated ICD-10-CM diagnosis codes or ICD-10-PCS procedure codes recorded during the index ablation hospitalization.

Acute kidney injury, acute stroke or TIA (either ischemic or hemorrhagic), complete or high-degree atrioventricular block, cardiac arrest, pulmonary embolism, mechanical circulatory support use, cardiogenic shock, cardiac tamponade and pericardial effusion without tamponade.

Major bleeding was coded to evaluate hemorrhage at multiple anatomical sites: postprocedural hemorrhage of the circulatory system, retroperitoneal hemorrhage, hemoperitoneum, hemothorax, hemopericardium, acute posthemorrhagic anemia, intracranial hemorrhage and gastrointestinal hemorrhage (hematemesis, melena, unspecified GI hemorrhage).

Vascular and access-site complications were defined using a composite code set capturing the specific mechanisms of catheter-access vascular injury relevant to the transfemoral venous approach used in AF ablation. We included: postprocedural hematoma, acquired arteriovenous fistula, pseudoaneurysm of lower extremity artery, vascular device complication, direct femoral vein injury, femoral vein thrombosis attributable to access-site deep vein thrombosis, iliac vein thrombosis from high femoral puncture or sheath-related injury, and catheter or sheath-related vascular thrombosis

### Composite Endpoints

We evaluated two composite endpoints which we applied across the component binary indicators, such that a hospitalization was classified as a composite event if any one or more of the components was present.

Composite major adverse hospital event was defined as the occurrence of any of the following: in-hospital mortality, acute kidney injury, acute stroke or TIA, major bleeding, cardiogenic shock, pulmonary embolism or cardiac arrest.

Composite procedural complication was defined as the occurrence of any of the following procedure-specific adverse events: cardiac tamponade, pericardial effusion, vascular or access-site complication, or AV block.

### Covariate Identification

Clinical, demographic and administrative covariates were identified a priori. We included - age at admission, sex, race, median household income, primary expected payer, hospital location and teaching status, and hospital bed size.

Clinical comorbidities and their ICD-10-CM code definitions were as follows: Diabetes mellitus, Hypertension, Heart failure, Chronic kidney disease, Chronic obstructive pulmonary disease (COPD), Obesity, Dyslipidemia, Obstructive sleep apnea, Coronary artery disease, Prior myocardial infarction, Prior stroke or TIA, Smoking or tobacco use, Alcohol use disorder, Coagulopathy, Depression, Valvular heart disease, and Thyroid disease.

### Statistical Analysis

A propensity score (PS) for prevalent PAD was estimated using logistic regression on the full cohort. Our model included covariates identified a priori and included as predictors: age, sex, household income quartile, primary payer, AF subtype, hospital teaching status, hospital bed size, and 17 binary comorbidity indicators (hypertension, diabetes, heart failure, CKD, COPD, obesity, dyslipidemia, OSA, CAD, prior MI, prior stroke/TIA, smoking/tobacco use or nicotine dependence, alcohol use, coagulopathy, depression, valvular disease, thyroid disease). Calendar year was included as a categorical variable to account for temporal trends in PAD coding and ablation volume.

Model discrimination was assessed using the C-statistic area under the ROC curve derived from the propensity score.

Stabilized inverse probability of treatment weights (IPTW) were computed for each observation using the marginal probabilities of treatment group membership as the stabilization numerators.

This approach creates a pseudo-population in which PAD and non-PAD patients have similar covariate distributions, enabling estimation of the marginal average treatment effect.

We assessed covariate balance before and after weighting using standardized mean differences (SMD). A threshold of SMD <0.10 was used to define adequate covariate balance.

### Primary Outcome Analysis

Binary outcomes were analyzed using weighted logistic regression. Results are reported as adjusted odds ratios (aOR) with 95% empirical confidence intervals and two-sided p-values. The significance threshold was two-sided alpha = 0.05.

### Resource Utilization

Length of stay was analyzed using a negative binomial regression model. Results are reported as incidence rate ratios (IRR) with 95% confidence intervals. Total hospital charges were analyzed using a weighted gamma regression model. Results are reported as charge ratios with 95% confidence intervals.

### Sensitivity Analysis

A pre-specified sensitivity analysis was performed using conventional multivariable logistic regression on the full unweighted cohort. This approach avoids reliance on the IPTW pseudo-population and weight construction. All covariates used in the propensity score model were included simultaneously as adjustors. Models were fit separately for each outcome. The PAD indicator was coded as a binary class variable with the non-PAD group as the reference. Odds ratios, 95% Wald confidence intervals, and p-values for the PAD main effect are reported.

All statistical analyses were performed using SAS version 9.4 (SAS Institute, Cary, NC). Statistical significance was defined as two-sided p<0.05 throughout.

## RESULTS

### Study Cohort and PAD Prevalence

Our final sample comprised 22,166 AF catheter ablation hospitalizations of which 899 (4.06%) carried a concurrent diagnosis of peripheral artery disease and 21,267 (95.94%) did not.

### Baseline Characteristics

Patients with PAD who underwent AF ablation were on average 4.5 years older than those without (mean age 71.8 vs. 67.3 years; SD 9.67 vs. 11.94; p<0.001) and were more likely to be male (65.2% vs. 60.8%; p = 0.009) (Table 1). White patients made up 84.8% of the PAD group compared with 81.3% of the non-PAD group (p<0.001). PAD patients were within the lowest income quartile (30.1% vs. 25.2%; p = 0.0002) and were substantially more likely to hold Medicare as their primary payer (79.9% vs. 63.6%; p<0.001).

**Table 1.** Baseline Characteristics of AF Ablation Hospitalizations by PAD Status.

| Characteristic | No PAD<br>(n=21,267) | PAD (n=899) | p-value |
| --- | --- | --- | --- |
| **Demographics** |  |  |  |
| Age, mean years (SD) | 67.3 (11.94) | 71.8 (9.67) | **<0.001** |
| Age, median years [IQR] | 68 [60-76] | 72 [65-78] | **<0.001** |
| **Sex** |  |  |  |
| Male, % | 60.8 | 65.2 | **0.009** |
| Female, % | 39.2 | 34.8 |  |
| **Race/ethnicity** |  |  |  |
| White, % | 81.3 | 84.8 | **<0.001** |
| Black, % | 10.5 | 11.6 |  |
| Hispanic, % | 6.5 | 3.2 |  |
| Asian/other, % | 1.7 | 0.4 |  |
| **Socioeconomic and insurance** |  |  |  |
| Lowest income quartile (Q1), % | 25.2 | 30.1 | **<0.001** |
| Medicare primary payer, % | 63.6 | 79.9 | **<0.001** |
| **AF subtype (overall chi-square $p=0.019$)** |  |  |  |
| Paroxysmal, % | 58.7 | 60.0 |  |
| Persistent, % | 11.9 | 10.3 |  |
| Permanent, % | 2.6 | 4.1 |  |
| Unspecified, % | 26.8 | 25.6 |  |
| **Comorbidities** |  |  |  |
| Hypertension, % | 79.1 | 90.2 | **<0.001** |
| Coronary artery disease, % | 43.4 | 73.2 | **<0.001** |
| Dyslipidemia, % | 55.0 | 69.6 | **<0.001** |
| Diabetes mellitus, % | 31.1 | 25.0 | **<0.001** |
| Heart failure, % | 47.8 | 62.5 | **<0.001** |
| Chronic kidney disease, % | 23.4 | 37.4 | **<0.001** |
| COPD, % | 20.3 | 38.9 | **<0.001** |
| Obesity, % | 25.6 | 20.5 | **0.001** |
| Obstructive sleep apnea, % | 21.6 | 18.6 | **0.03** |
| Prior myocardial infarction, % | 11.0 | 22.0 | **<0.001** |
| Prior stroke or TIA, % | 8.4 | 13.7 | **<0.001** |
| Smoking or tobacco use, % | 39.8 | 56.5 | **<0.001** |
| Coagulopathy, % | 6.7 | 9.3 | **0.002** |
| Valvular heart disease, % | 11.9 | 14.8 | **0.008** |
| Alcohol use disorder, % | 3.1 | 2.8 | 0.58 |
| Depression, % | 0.5 | 0.7 | 0.47 |
| Thyroid disease, % | 1.4 | 1.2 | 0.73 |
| **Hospital characteristics** |  |  |  |
| Urban teaching hospital, % | 86.1 | 83.0 | **0.01** |
| Large-bed hospital, % | 63.4 | 59.4 | **0.02** |
| **Resource utilization (unweighted)** |  |  |  |
| LOS, mean days (SD) | 5.2 (6.35) | 6.4 (8.16) | **<0.001** |
| LOS, median days [IQR] | 3 [2-6] | 5 [3-8] | **<0.001** |
| Total charges, mean (SD) | \$166,897<br>(\$139,348) | \$188,078<br>(\$170,570) | **<0.001** |

*AF, atrial fibrillation; COPD, chronic obstructive pulmonary disease; IQR, interquartile range; LOS, length of stay; PAD, peripheral artery disease; SD, standard deviation; TIA, transient ischemic attack. p-values from Pearson chi-square for categorical variables and independent samples t-test for continuous variables. Bold p-values indicate $p < 0.05$ .*

Atherosclerotic risk factor burden was uniformly higher among PAD patients. Coronary artery disease was present in 73.2% of PAD patients versus 43.4% of those without PAD (p<0.001). Hypertension (90.2% vs. 79.1%), COPD (38.9% vs. 20.3%), dyslipidemia (69.6% vs. 55.0%), chronic kidney disease (37.4% vs. 23.4%), prior myocardial infarction (22.0% vs. 11.0%), prior stroke or TIA (13.7% vs. 8.4%), and smoking or tobacco use (56.5% vs. 39.8%) were all significantly more common in the PAD group (all p<0.001). Coagulopathy (9.3% vs. 6.7%; p=0.002) and valvular heart disease (14.8% vs. 11.9%; p=0.008) were also more prevalent. Diabetes mellitus was paradoxically less common among PAD patients (25.0% vs. 31.1%; p <0.001), as was obesity (20.5% vs. 25.6%; p=0.001), a pattern consistent with survival selection bias in this ablation-selected population.

The AF subtype distribution differed significantly between groups (chi-square p=0.019). Paroxysmal AF was the most common subtype in both (60% in PAD vs. 58.7% in No PAD), but permanent AF was proportionally more frequent among PAD patients (4.1% vs. 2.6%), suggesting a more advanced atrial disease process. Before weighting, PAD patients had longer mean hospital stays (6.4 vs. 5.2 days; p<0.001) and higher mean total charges ($186,093 vs. $166,937; p<0.001).

### Propensity Score Model

The logistic propensity score model for PAD, built on prespecified demographic, clinical, and institutional covariates, demonstrated good discrimination with a C-statistic of 0.76 (95% CI 0.74 to 0.77). Among the strongest positive predictors of prevalent PAD were coronary artery disease (OR 2.43; 95% CI, 2.05 - 2.88), COPD (OR 1.70; 95% CI, 1.46 - 1.98), smoking/tobacco exposure (OR 1.54; 95% CI, 1.33 - 1.78), hypertension (OR 1.49; 95% CI, 1.18 - 1.89), chronic kidney disease (OR 1.41; 95% CI, 1.21 - 1.64), dyslipidemia (OR 1.35; 95% CI, 1.15 - 1.57), prior stroke or TIA (OR 1.32; 95% CI, 1.08 - 1.62), and older age (OR 1.02 per year; 95% CI, 1.01 - 1.03). Diabetes mellitus was inversely associated with PAD (OR 0.51; 95% CI 0.44 - 0.60), likely reflecting the complex drawbacks of administrative data and coding.

### Covariate Balance

Covariate balance before and after weighting is shown in Table 2. Seventeen of 19 prespecified covariates achieved the post-weighting balance threshold of SMD <0.10. Residual imbalance remained for age although both were improved compared with the pre-weighting cohort.

**Table 2.** Standardized Mean Differences (SMD) Before and After Stabilized IPTW Weighting.

| Covariate | Pre-IPTW SMD | Post-IPTW SMD |
| --- | --- | --- |
| **Age (years)** | 0.418 | **0.174** |
| Coronary artery disease | 0.634 | 0.031 |
| COPD | 0.417 | 0.044 |
| Hypertension | 0.312 | 0.067 |
| Chronic kidney disease | 0.307 | 0.087 |
| Dyslipidemia | 0.305 | 0.059 |
| **Heart failure** | 0.298 | **0.104** |
| Prior myocardial infarction | 0.299 | 0.056 |
| Smoking or tobacco use | 0.340 | 0.011 |
| Diabetes mellitus | 0.136 | 0.080 |
| Prior stroke or TIA | 0.170 | 0.018 |
| Obesity | 0.122 | 0.056 |
| Female sex | 0.090 | 0.013 |
| Coagulopathy | 0.099 | 0.003 |
| Obstructive sleep apnea | 0.074 | 0.023 |
| Valvular heart disease | 0.086 | 0.015 |
| Alcohol use disorder | 0.019 | 0.057 |
| Depression | 0.023 | 0.011 |
| Thyroid disease | 0.012 | 0.028 |

*SMD, standardized mean difference. The prespecified balance target was SMD <0.10. Residual imbalance persisted for age and heart failure*

### Primary Outcome: In-Hospital Mortality

In-hospital mortality was observed in a weighted equivalent of 11 PAD patients (1.3%) and 225 non-PAD patients (1.1%) corresponding to an adjusted odds ratio of 1.32 (95% CI, 0.66 - 2.64; p = 0.44) in our regression model. In-hospital mortality was not significantly associated with PAD in this analysis.

While the point estimate is consistent with a mortality excess in PAD patients, the data does not meet statistical significance and the wide confidence interval reflects the small absolute number of deaths attributable to PAD patients.

### Secondary Outcomes

Results for all pre-specified secondary and composite outcomes are shown in Table 3.

**Table 3.** Primary and Secondary Outcomes (PAD vs. No PAD; n=22,166)

| **Outcome** | **PAD %**<br>**(n)** | **No**<br>**PAD %**<br>**(n)** | **aOR** | **95% CI** | **p-value** |
| --- | --- | --- | --- | --- | --- |
| **Primary outcome** |  |  |  |  |  |
| In-hospital mortality | 1.39<br>(12) | 1.06<br>(221) | 1.32 | 0.66-2.64 | 0.44 |
| **Secondary outcomes** |  |  |  |  |  |
| **Major bleeding** | 9.17<br>(79) | 5.87<br>(1,242) | **1.62** | 1.17-2.24 | **<0.01** |
| **Vascular/access-site complication** | 2.52<br>(22) | 1.42<br>(300) | **1.80** | 1.04-3.12 | **0.04** |
| **Acute kidney injury** | 22.69<br>(195) | 18.28<br>(3,872) | **1.31** | 1.05-1.64 | **0.02** |
| Acute stroke or TIA | 1.26<br>(11) | 1.40<br>(300) | 0.90 | 0.47-1.72 | 0.76 |
| Cardiogenic shock | 3.76<br>(32) | 3.14<br>(669) | 1.20 | 0.74-1.97 | 0.46 |
| Cardiac tamponade | 2.63<br>(23) | 1.75<br>(369) | 1.51 | 0.78-2.94 | 0.22 |
| Pericardial effusion | 5.50<br>(47) | 5.68<br>(1,208) | 0.97 | 0.65-1.45 | 0.87 |
| Mechanical circulatory support | 1.52<br>(13) | 1.14<br>(246) | 1.35 | 0.79-2.28 | 0.27 |
| AV block | 7.10<br>(61) | 6.73<br>(1,426) | 1.06 | 0.66-1.70 | 0.81 |
| Cardiac arrest | 1.50<br>(13) | 0.84<br>(179) | 1.80 | 0.92-3.54 | 0.09 |
| Pulmonary embolism | 0.40 (4) | 0.50<br>(108) | 0.81 | 0.30-2.16 | 0.67 |
| **Composite endpoints** |  |  |  |  |  |
| **Composite major adverse hospital event** | 29.42<br>(254) | 24.41<br>(5,183) | **1.29** | 1.05-1.59 | **0.02** |
| Composite procedural complication | 12.14<br>(105) | 9.65<br>(2,043) | 1.29 | 0.92-1.82 | 0.14 |

**Table 4.** Hospital Resource Utilization in the IPTW-Weighted Analysis.

| **Measure** | **PAD (n=899)** | **No PAD (n=21,267)** | **Estimate (95% CI)** | **p-value** |
| --- | --- | --- | --- | --- |
| LOS, mean days (SD) | 6.4 (8.16) | 5.2 (6.35) | IRR 1.14<br>(0.95-1.36) | 0.17 |
| Total charges, mean (SD) | \$188,078<br>(\$170,570) | \$166,897<br>(\$139,348) | **Ratio 1.13**<br>**(1.04-1.22)** | **0.003** |

*IRR, incidence rate ratio. 95% CI, empirical Wald confidence interval. Bold estimates indicate statistical significance at two-sided alpha 0.05.*

Major bleeding was significantly more common in PAD hospitalizations (9.17% vs 5.87%; aOR 1.62; 95% CI, 1.17 - 2.24; p = 0.004), as did vascular/access-site complications (2.52% vs 1.42%; aOR 1.80; 95% CI, 1.04 - 3.12; p = 0.04) and acute kidney injury (22.69% vs 18.28%; aOR 1.31; 95% CI, 1.05 - 1.64; p = 0.02).

Among remaining secondary outcomes, none reached statistical significance. Cardiac arrest occurred in 1.50% of PAD patients and 0.84% of non-PAD patients (aOR 1.80; 95% CI 0.92 - 3.54; p = 0.09). Cardiac tamponade was numerically higher in PAD (2.63% vs. 1.75%; aOR 1.51; 95% CI 0.78 - 2.94; p= 0.22). Mechanical circulatory support use was 1.52% in PAD versus 1.14% in non-PAD patients (aOR 1.35; 95% CI 0.79 to 2.28; p = 0.27). Acute stroke or TIA occurred at similar rates (1.26% vs. 1.40%; aOR 0.90; 95% CI 0.47 to 1.72; p = 0.76). Pericardial effusion was also comparable across groups (5.50% vs. 5.68%; aOR = 0.97; CI = 0.65 - 1.45; p = 0.87). Cardiogenic shock, pulmonary embolism, and the composite procedural complication endpoint were also non-significant.

Composite major adverse hospital event (mortality, AKI, stroke, major bleeding, cardiogenic shock, pulmonary embolism, and cardiac arrest) occurred in 29.42% of PAD patients versus 24.41% of non-PAD patients (aOR 1.29; 95% CI, 1.05 - 1.59; p = 0.02). Composite procedural complication (tamponade, pericardial effusion, vascular complication, and AV block) occurred in 12.14% of PAD patients versus 9.65% of non-PAD patients (aOR 1.29; 95% CI 0.92 - 1.82; p = 0.14), which did not reach statistical significance.

### Hospital Resource Utilization

Mean length of stay in the final cohort was 6.4 days in PAD patients and 5.2 days in non-PAD patients. Weighted negative binomial regression yielded an incidence rate ratio of 1.14 (95% CI, 0.95 - 1.36; p = 0.17), indicating no statistically significant difference in adjusted length of stay. Mean total hospital charges were $188,078 in PAD hospitalizations and $166,897 in non-PAD hospitalizations. Weighted gamma regression yielded a charge ratio of 1.13 (95% CI, 1.04 - 1.22; p = 0.003), indicating approximately 13% higher adjusted charges in the PAD group.

### Sensitivity Analysis: Multivariable Logistic Regression

In the prespecified multivariable sensitivity analysis (table 5), major bleeding remained significantly elevated in PAD hospitalizations (aOR 1.34; 95% CI, 1.05 - 1.73; p = 0.02), as did vascular/access-site complications (aOR 1.63; 95% CI, 1.03 - 2.59; p = 0.04), cardiac arrest (aOR 1.76; 95% CI, 1.00 - 3.10; p = 0.049), and composite major adverse hospital event (aOR 1.19; 95% CI, 1.02 - 1.39; p = 0.03). In-hospital mortality remained non-significant (aOR 0.96; 95% CI, 0.53 - 1.76; p = 0.90). Acute kidney injury (aOR 1.16; 95% CI, 0.98 - 1.38; p = 0.09) and mechanical circulatory support (aOR 1.53; 95% CI, 0.95 - 2.48; p=0.08) remained borderline.

**Table 5.** Sensitivity Analysis: Multivariable-Adjusted Odds Ratios for PAD vs. No PAD.

| Outcome | aOR | 95% CI | p-value |
| --- | --- | --- | --- |
| **Primary outcome** |  |  |  |
| In-hospital mortality | 0.96 | 0.53-1.76 | 0.90 |
| **Secondary outcomes** |  |  |  |
| Major bleeding | 1.34 | 1.05-1.73 | **0.02** |
| Vascular/access-site complication | 1.63 | 1.03-2.59 | **0.04** |
| Cardiac arrest | 1.76 | 1.00-3.10 | **0.049** |
| Acute kidney injury | 1.16 | 0.98-1.38 | 0.09 |
| Acute stroke or TIA | 1.04 | 0.59-1.84 | 0.90 |
| Cardiogenic shock | 1.13 | 0.79-1.61 | 0.52 |
| Cardiac tamponade | 1.28 | 0.76-2.16 | 0.35 |
| Pericardial effusion | 0.95 | 0.69-1.32 | 0.76 |
| Mechanical circulatory support | 1.53 | 0.95-2.48 | 0.08 |
| AV block | 0.92 | 0.70-1.19 | 0.51 |
| Pulmonary embolism | 1.08 | 0.43-2.71 | 0.87 |
| **Composite endpoints** |  |  |  |
| Composite major adverse hospital event | 1.19 | 1.02-1.39 | **0.03** |
| Composite procedural complication | 1.08 | 0.87-1.34 | 0.46 |

All remaining outcomes were non-significant. These findings corroborate the primary propensity weighted analysis with respect to major bleeding and vascular complications

## DISCUSSION

In this nationwide analysis of adults undergoing inpatient AF catheter ablation, PAD was associated with higher periprocedural morbidity but not with a statistically significant increase in in-hospital mortality. The clearest signals were major bleeding and vascular or access-site complications, and both remained significant in the prespecified multivariable sensitivity analysis. AKI and composite major adverse hospital event were also higher in the primary analysis, although the AKI association attenuated in the sensitivity model. PAD was not significantly associated with stroke or TIA, cardiac tamponade, pericardial effusion, pulmonary embolism, or complete atrioventricular block. Because deaths were infrequent in the PAD group, the no difference in mortality finding should be interpreted cautiously rather than as evidence of equivalence.

Several mechanisms may explain these associations. PAD reflects systemic atherosclerosis, endothelial dysfunction, vascular calcification, and high comorbidity burden.^14–16^ In the setting of transfemoral venous access, vascular calcification and altered anatomic landmarks may increase the risk of difficult access, inadvertent arterial puncture, hematoma, pseudoaneurysm, or access-site thrombosis. PAD patients in our cohort also had substantially higher rates of coronary artery disease, chronic kidney disease, heart failure, COPD, prior myocardial infarction, and coagulopathy. These comorbidities may amplify periprocedural risks, particularly when combined with the systemic anticoagulation required for AF ablation and potential antiplatelet exposure in patients with established vascular disease.^19^ The observed increase in AKI may reflect reduced renal reserve and hemodynamic susceptibility rather than a direct PAD-specific effect, consistent with its attenuation after adjustment for CKD.

Resource utilization findings further highlight the clinical impact of these associations. Although length of stay was not significantly different between PAD and non-PAD patients undergoing AF ablation, the total hospital charges remained higher in PAD patients. This suggests that comorbid PAD may be associated with increased healthcare utilization, which may reflect greater post-procedural monitoring, diagnostic evaluation, or management of complications such as bleeding, vascular injury, or renal dysfunction.

Our findings extend the broader AF literature showing that PAD identifies patients with higher long-term cardiovascular and bleeding risk.^17,18^ Direct evidence in the ablation setting remains sparse. In a single-center study of AF ablation, peripheral vascular disease was independently associated with early cerebral thromboembolic complications,^20^ but bleeding, access-site complications, AKI, and resource use were not assessed in the outcomes. By evaluating PAD as a distinct exposure in a large, nationally representative inpatient AF ablation cohort, our study adds clinically relevant data showing that PAD is associated with greater procedural morbidity, especially bleeding and vascular or access-site complications.

These findings have practical implications. PAD status may be worth incorporating into pre-ablation risk assessment and counseling, especially when discussing bleeding risk, vascular access complications, renal vulnerability, and expected resource use. Current AF ablation guidelines acknowledge the importance of overall comorbidity burden, but they do not identify PAD specifically as a procedural risk modifier.^3,5^ The higher risk of access-site and vascular complication is notably important given the ULYSSES randomized trial, ^25^ which showed fewer access-site complications with ultrasound-guided venous access than with conventional puncture. While the trial did not stratify by PAD, the findings may define a subgroup with greater absolute benefit from ultrasound-guided access. This hypothesis needs prospective testing. The increased risk of AKI also supports careful renal-protective planning, including hydration, contrast minimization when feasible, and avoidance of additional nephrotoxins.

Future studies should validate these findings in procedural registries with granular clinical and procedural detail, including PAD severity, prior peripheral revascularization, antithrombotic therapy, access strategy, sheath size, closure technique, ablation modality, operator experience, and outpatient outcomes. Several literature gaps remain. We do not know whether risk differs by symptomatic vs asymptomatic PAD, by prior limb revascularization, by combined anticoagulant-antiplatelet exposure, or by modern workflow choices such as ultrasound-guided access and same-day discharge. It also remains unclear whether newer ablation technologies, including pulsed field ablation, which has shown favorable overall safety in contemporary studies,^10–12^ modify complication risk in patients with PAD.

### Limitations

This study has several limitations inherent to administrative database research. The NIS is a discharge-level database, not a patient-level procedural registry; therefore, repeated hospitalizations cannot be linked, longitudinal outcomes cannot be assessed, and arrhythmia recurrence, post-discharge complications, and readmissions are unavailable. Because the NIS captures inpatient hospitalizations only, our cohort likely represents a higher-risk subset of AF ablation procedures and may not generalize to contemporary outpatient or same-day-discharge practice.

Exposure and outcome definitions relied on ICD-10-CM and ICD-10-PCS codes and are therefore vulnerable to misclassification. Contemporary ICD-10-PCS codes are not fully specific for pulmonary vein isolation, as shown by Ferro et al,^23^ and our pacemaker-based exclusion rule may not have eliminated all atrioventricular junction ablation cases. PAD coding in administrative data may also under detect milder or asymptomatic disease, and claims data do not capture PAD severity, symptomatic status, prior peripheral revascularization, anatomic distribution, or the burden of iliofemoral calcification.

Administrative coding may not reliably distinguish pre-existing diagnoses from incident postprocedural complications for all outcomes, particularly acute kidney injury, pericardial effusion, atrioventricular block, and vascular complications. Medication and procedural details were unavailable, including anticoagulant and antiplatelet therapy, intraprocedural anticoagulation strategy, access technique, sheath size, closure method, contrast use, operator volume, and ablation complexity. We also could not identify ablation energy modality and therefore could not assess whether ablation technology modifies procedural risk in PAD patients.

Although stabilized IPTW improved balance across most measured covariates, residual imbalance and unmeasured confounding from frailty, functional status, PAD severity, medication use, and center-level practices cannot be excluded.

The small number of deaths among PAD hospitalizations limited power for the mortality end point, so the nonsignificant result should not be interpreted as equivalence. Some outcome categories may also overlap, particularly bleeding and access-site complications. Accordingly, the composite end points should be interpreted as summary measures of in-hospital adverse events rather than as distinct outcomes.

## Conclusion

Among adults undergoing inpatient AF catheter ablation in the United States from 2016 through 2020, PAD was associated with higher odds of major bleeding, vascular or access-site complications, acute kidney injury, composite major adverse hospital events, and higher hospital charges, without significant difference in in-hospital mortality. The most consistent outcomes were bleeding and vascular complications across both analyses. These data support routine assessment of PAD status before AF ablation, careful individualized antithrombotic, anticoagulant and renal-protective planning, and prospective study of whether contemporary access strategies and newer ablation technologies can mitigate risk in this population.

## Data Availability

Data is publicly available by HCUP/AHRQ.

## ACKNOWLEDGMENTS

None.

## SUPPLEMENTAL MATERIAL

Tables S1 - S12 (ICD-10-CM and ICD-10-PCS Code Definitions)

## Non-standard Abbreviations and Acronyms

AF: atrial fibrillation
AKI: acute kidney injury
aOR: adjusted odds ratio
AV: atrioventricular
CAD: coronary artery disease
CI: confidence interval
CKD: chronic kidney disease
COPD: chronic obstructive pulmonary disease
IPTW: inverse probability of treatment weighting
IRR: incidence rate ratio
LOS: length of stay
MCS: mechanical circulatory support
NIS: National Inpatient Sample
OSA: obstructive sleep apnea
PAD: peripheral artery disease
PFA: pulsed field ablation
PS: propensity score
PVI: pulmonary vein isolation
SD: standard deviation
SMD: standardized mean difference
TIA: transient ischemic attack.

